# Built to last? Reproducibility and Reusability of Deep Learning Algorithms in Computational Pathology

**DOI:** 10.1101/2022.05.15.22275108

**Authors:** Sophia J. Wagner, Christian Matek, Sayedali Shetab Boushehri, Melanie Boxberg, Lorenz Lamm, Ario Sadafi, Dominik J. E. Waibel, Carsten Marr, Tingying Peng

## Abstract

Recent progress in computational pathology has been driven by deep learning. While code and data availability are essential to reproduce findings from preceding publications, ensuring a deep learning model’s reusability is more challenging. For that, the codebase should be well-documented and easy to integrate in existing workflows, and models should be robust towards noise and generalizable towards data from different sources. Strikingly, only a few computational pathology algorithms have been reused by other researchers so far, let alone employed in a clinical setting.

To assess the current state of reproducibility and reusability of computational pathology algorithms, we evaluated peer-reviewed articles available in Pubmed, published between January 2019 and March 2021, in five use cases: stain normalization, tissue type segmentation, evaluation of cell-level features, genetic alteration prediction, and direct extraction of grading, staging, and prognostic information. We compiled criteria for data and code availability, and for statistical result analysis and assessed them in 161 publications. We found that only one quarter (42 out of 161 publications) made code publicly available and thus fulfilled our minimum requirement for reproducibility and reusability. Among these 42 papers, three quarters (30 out of 42) analyzed their results statistically, less than half (20 out of 42) have released their trained model weights, and only about a third (16 out of 42) used an independent cohort for evaluation.

This review highlights candidates for reproducible and reusable algorithms in computational pathology. It is intended for both pathologists interested in deep learning, and researchers applying deep learning algorithms to computational pathology challenges. We provide a list of reusable data handling tools and a detailed overview of the publications together with our criteria for reproducibility and reusability.

## 1 Introduction

Technical progress has been driving digitization in pathology over the past decade. Coupled with advances in deep learning methods, computational approaches help to localize, segment, and classify single cells and tissue types in an automated manner - and form the research field of computational pathology (see Box 1 for a glossary; (Fuchs and Buhmann, 2011). In particular, deep neural networks have recently been shown to reach the performance level of medical experts on well-defined tasks such as skin cancer diagnosis (Esteva et al., 2017), lung cancer subtype classification (Coudray et al., 2018), or the recognition of malignant white blood cells (Matek et al., 2019)

However, despite the steady increase in the number of publications in this field (Fig. 1) and their promising results, only a few have reached clinical implementation (Echle et al., 2020a; van der Laak et al., 2021). This is due to several reasons: For deep learning-based methods, code availability is a natural requirement for reproducibility, which, unfortunately, is not yet current practice for most publications. Even when code is available, reproducing the original results can be challenging and requires the assistance of the original author (Pineau et al., 2020). In particular, ready-to-use scripts with sufficient instructions or intuitive demo examples are rarely published. This makes the reuse of recent methods difficult for non-deep-learning experts, especially for pathologists who are not supported by computational experts. Another reason, which is particularly relevant to clinical implementation, is the generalization gap of algorithms in computational pathology. Often, the published performance of deep learning algorithms cannot be transferred to other datasets, due to differences in staining or scanner settings. Therefore, external validation of algorithms and statistical robustness analysis are key to assess generalizability. Finally, any algorithm employed in a clinical setting must additionally be approved by national or international authorities such as the United States Food and Drugs Administrative (FDA) or the European Medicines Agency (EMA), an often lengthy and complicated process involving business markets and legal issues, which is beyond the scope of this review.

**Figure 1:**
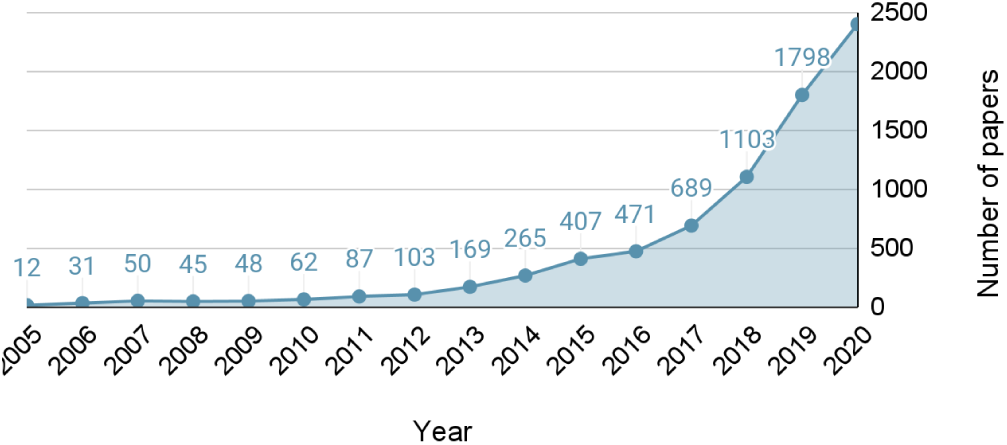
The number of papers published in the field of computational pathology in PubMed [retrieved on 22nd July ‘21] has markedly increased in the last 15 years.

Here, we focus on deep learning algorithms for computational pathology and their reproducibility and reusability. For the ultimate goal of *reusing* other deep learning algorithms, the algorithms must be *reproducible* and generalizable to similar datasets (i.e., *robust*) and external datasets (i.e., *replicable*) (see Box 2 and Fig. 2 for a definition of reproducibility and reusability and related terms). Because hematoxylin-and-eosin (H&E) staining is the most commonly used routine staining for cancer diagnosis and is often referred to as the baseline staining (Rosai, 2007), we restrict this review to methods for H&E-stained whole-slide image (WSI) analysis. We considered five computational pathology use cases and assembled a systematic overview of publications published between January 2019 and March 2021. For this, we compiled criteria for reproducibility in a practical context and examined each work with respect to these. We additionally provide an overview of current data handling tools.

**Figure 2:**
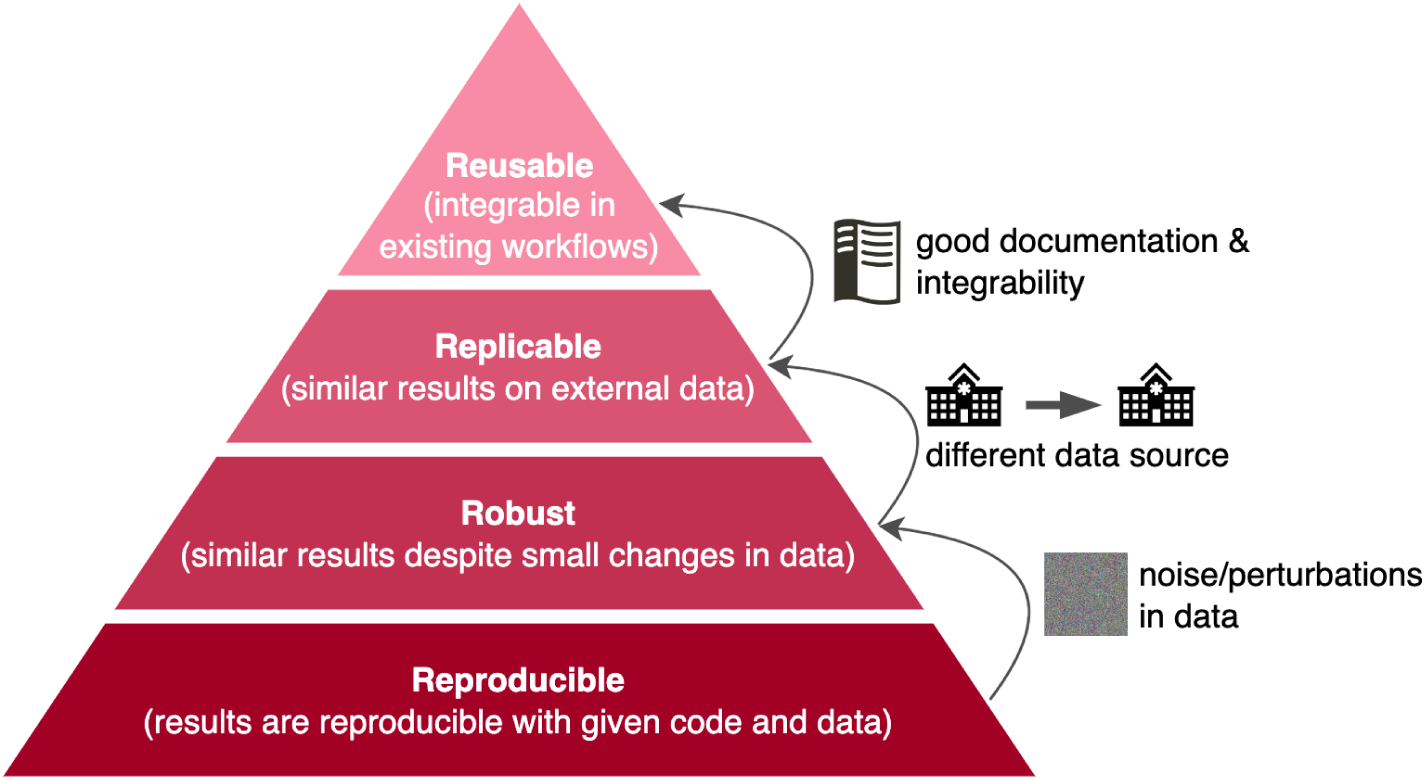
Reproducibility, robustness, replicability, and reusability in the context of deep learning algorithms for computational pathology.

### Box 1

**Glossary**

#### Digital pathology

Histological slides are scanned and digitized, such that pathologists can examine the patient material on a computer instead of working on optical microscopes. Digitized slides can be stored and processed, enabling the use of computational methods in the diagnostic process.

#### Computational pathology

The analysis of digitized histological slides with computational methods (Fuchs and Buhmann, 2011).

#### Specimen

A tissue sample, e.g., obtained during a biopsy or other surgical procedures, typically fixed in formalin and embedded in paraffin (FFPE).

#### Section

A thin slice (with a typical thickness of 3-15 µm) of a specimen mounted on a microscopic slide.

#### Whole-slide image (WSI)

The digitized image of a tissue section on a microscope slide. Slides can be scanned in very high magnification resulting in images of sizes up to several giga-pixels.

#### Tiles and patches

WSIs are split up into smaller images (e.g. 512×512 pixels), also called patches, that can be processed by neural networks. If the patches are used for subsequent WSI-rendering by stitching them back together, they are often called tiles. Unlike WSIs, these smaller units of image data allow for easier and parallelized image processing.

#### Annotations

Diagnostic information on pixel-level or patch-level are obtained from manual expert pathologist labeling. WSI-level annotations can be all diagnostic information about the patient (e.g., age, survival, staging, grading) mostly obtained without additional expert pathologist interaction.

#### Supervised learning

Training procedure of a neural network, where the ground truth, i.e., the correct label for the task, is available for each data point. However, in medical imaging and especially in computational pathology, full expert annotations at pixel-level or patch-level are very time-consuming and hence rare. Pixel-level annotations are used to localize tissues in **segmentation** tasks, where each pixel is assigned a tissue label. Patch-level annotations are used for **classification** tasks, where one label is predicted for the entire input patch.

#### Weakly supervised learning

Due to the rareness of fully annotated WSIs, weakly supervised learning approaches, like multiple-instance learning (MIL), are often used to train neural networks. With MIL only WSI-level annotations, such as diagnostic information on the cancer type or survival, are required for classification.

#### Convolutional neural network (CNN)

A neural network that can be trained to extract features by sliding learnable filters across the image. This makes CNNs translationally invariant and therefore well suited for histological data since important features can be found anywhere on a tile.

#### U-Net

A powerful CNN with an encoder-decoder architecture used for segmenting biomedical images (Ronneberger et al., 2015). It was adapted in many different ways and ranks among the most common architectures for segmentation tasks.

#### Mask R-CNN

A CNN architecture for instance segmentation in object detection (He et al., 2017). In contrast to U-Net that does not distinguish between instances of a class, Mask R-CNN outputs a segmentation mask for each instance on the image, which makes it useful for tasks such as nuclei segmentation and cell counting.

### Box 2

**Definitions**

#### Reproducibility

Using identical materials and procedures, the results of a study can be duplicated, and ultimately, identical conclusions can be drawn (Goodman et al., 2016). In the context of algorithms, the same result can be obtained from the same data, code, and analysis methods (Artner et al., 2020; Pineau et al., 2020).

#### Robustness

The same results are obtained from an algorithm despite small perturbations in the input (Li, 2018; Oala et al., 2020).

#### Replicability

Conclusions are stable based on independently acquired data (Artner et al., 2020; Pineau et al., 2020), i.e., code and analysis methods can be employed to external data with similar results and performance. For deep learning algorithms, replicability is equivalent to model generalizability, which is a key requirement for the clinical application of new algorithms.

#### Reusability

A piece of software is considered reusable if it can be included in an existing computational pathology setup with minor efforts (e.g., without the need for extensive rearrangements of the workflow).

## 2 Use Cases

We collected 161 papers from January 2019 and March 2021 on the automated analysis of histological slides for cancer diagnosis and treatment (Supplementary Table T1-5). We split this body of literature into the following use cases: (i) stain normalization, (ii) tissue type segmentation, (iii) evaluation of cell-level features, (iv) genetic alteration prediction, and (v) direct extraction of grading, staging, and prognostic information (Fig. 3).

**Figure 3:**
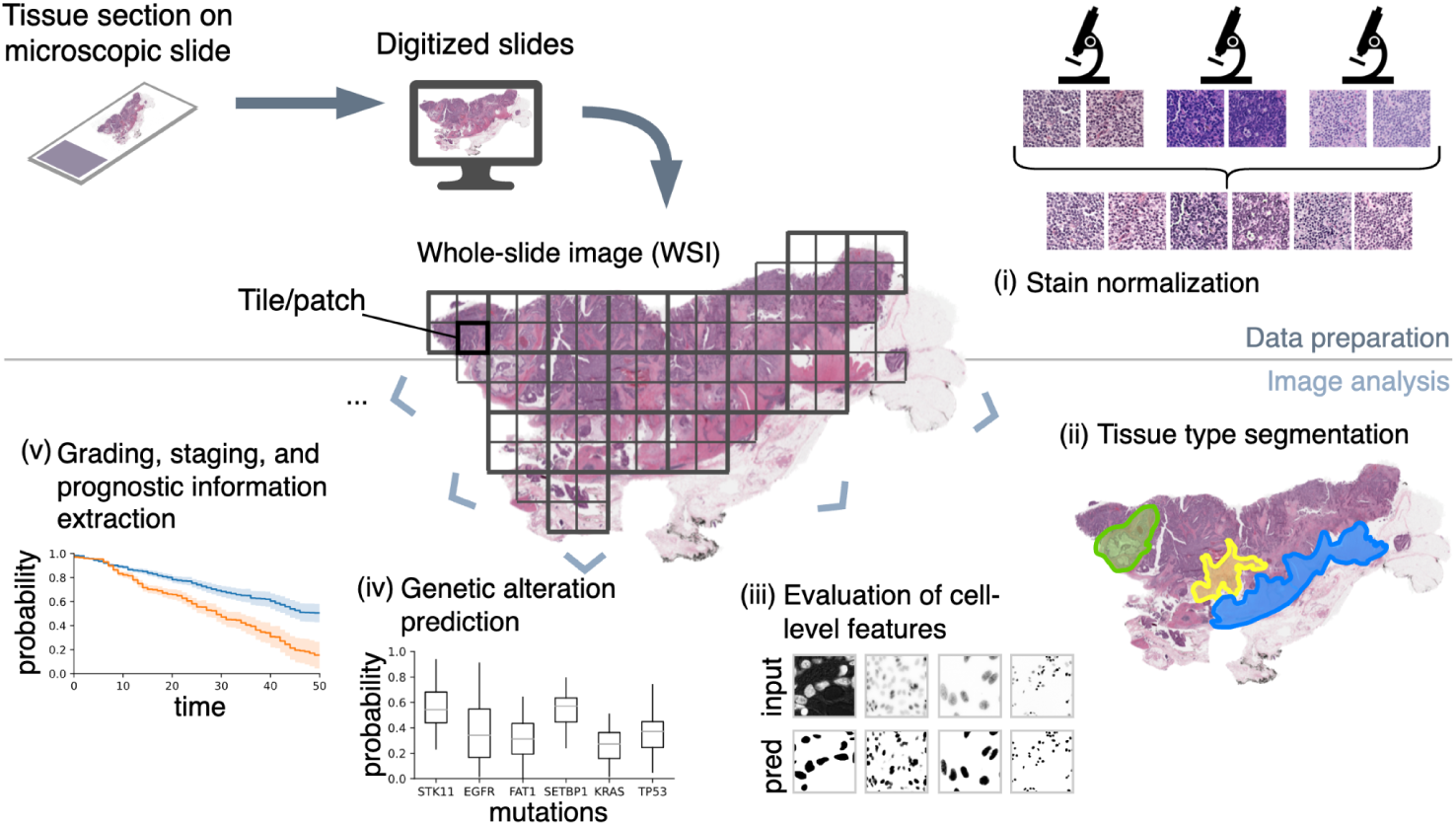
Overview of the use of deep learning in computational pathology including data handling tools for reading, annotating, and sharing WSIs (Box 3) and five applications of deep learning methods, which are covered in Section 2.1 - 2.5

In this technical chapter, we first provide a brief introduction of every use case, followed by an overview of the latest deep learning methods with a focus on works that provide code along with the publication. At the end of each section, we wrap up with an analysis of the reproducibility in the specific context.

As a prerequisite to any use case, open and publicly available data handling tools for reading, annotating, and sharing histopathological data are essential. We compiled the most common tools in Box 3 and provide a more detailed overview in the supplementary material (Supplementary Table T0) on software features, requirements, and the possibility to extend the tool with its own code.

### Box 3

**Data Handling Tools**

A key prerequisite for implementing, transferring, and reusing computational pathology algorithms between researchers and different labs or institutions is a software structure that allows for the exchange of image data, annotations, and meta-information. With the progress of computational pathology, numerous data handling tools have been developed. In many tools, image data handling is based on system-level libraries like OpenSlide (Goode et al., 2013) or OMERO (Allan et al., 2012). They enable data to be interoperable between different vendor-specific image formats. Most of these tools also provide user interfaces for pathologists to annotate images. Annotations can include class labeling or point flags, geometric shapes, and image-level labeling. While many popular image data handling and annotation tools were developed as standalone packages (e.g., SlideRunner (Aubreville et al., 2018), QuPath (Bankhead et al., 2017), ASAP^1^) an increasing number of recently developed packages, such as Cytomine (Marée et al., 2016) or EXACT (Marzahl et al., 2021) allow for web-based, collaborative data handling, which is essential for distributing, exchanging, and annotating data as well as evaluating models in a multi-institutional setting.

In addition to image annotation and exchange, data-handling packages allow integration with independently developed analysis algorithms at different levels. Some tools offer integrated scripting for automation of the tasks, e.g., using Groovy in QuPath. Additionally, programming interfaces to popular machine learning languages such as Python have been developed, e.g., for OMERO. Several tools, such as EXACT and CaMicroscope^2^, offer integrated, server-side evaluation of deep learning models. A detailed overview of open and publicly available data handling tools and their respective functionalities is provided in Supplementary Table T1.

### 2.1 Stain Normalization

Most work in computational histopathology focuses on H&E-stained routine sections, with hematoxylin staining nuclei in blue-purple and eosin staining extracellular material in pink (Chan, 2014). Digitized sections are prone to a multitude of image variations, caused by differences in the tissue preparation technique (e.g. the thickness and flatness of the sample cut), staining protocols, handling, and storage conditions. Moreover, slide scanners differ in microscope illumination, image post-processing, or noise handling. These factors lead to large variability in the visual appearance of WSIs that affects subsequent analysis and may lead to poor generalizability of algorithms. Computational methods aim at reducing the effects of these variations (Chen et al., 2017), e.g., by normalizing the stain color from a predefined source domain to one or more target domains, to arrive at a comparable visual appearance. These methods include improvements of analytical approaches, such as color deconvolution, and more recently, deep learning-based methods.

#### Color space methods

Color deconvolution separates the hematoxylin from the eosin component in optical density space based on a reference tile (Macenko et al., 2009). This approach has been developed further recently: Adaptive color deconvolution (Zheng et al., 2019) incorporated the underlying stain distribution of the target WSI instead of only a single tile. There, the authors assumed that each pixel is assigned to one stain and fitted a deconvolution matrix to a group of randomly sampled pixels from the source WSI using gradient-based optimization. Alternatively, non-negative matrix factorization has been used to obtain a color deconvolution matrix (Vahadane et al., 2016) and was optimized for GPU usage (Anand et al., 2019).

#### Generative models

The increasing popularity of Generative Adversarial Networks (GANs) has led to the development of style-transfer methods for stain normalization (Tschuchnig et al., 2020), which can be trained on all target WSI tiles instead of expert-picked reference tiles (Liang et al., 2020). StainGAN was trained with a cycle-consistency loss between the source and target domain, and the generator of the target domain was used to normalize all images in that domain (Shaban et al., 2019b). However, GANs may not always preserve the tissue structure (Cohen et al., 2018). To overcome this, StainNet trained a convolutional neural network (CNN) consisting of 1×1-convolutions and thereby transformed the source image from its original color space via intermediate color spaces to the target color space without losing structural information (Kang et al., 2020). However, this approach relies on image pairs from two different domains, which is challenging as paired images rarely exist. Alternatively, specific loss functions that compare images before and after normalization can be used to preserve the histopathological information including texture, structure, and color features added to the traditional GAN-loss that learns the stain distribution of a reference dataset (Liang et al., 2020). In contrast to normalization methods, GANs can also be used to simulate stain variability by generating synthetic images. This renders neural networks on downstream tasks more robust and avoids losing relevant information due to limitations of normalization methods. Yamashita et al. (2021) propose data augmentation based on style transfer from artistic paintings by replacing the low-level texture content with the style of artistic images. Since this produces unrealistic images for augmentation, Wagner et al. (2021) use a GAN architecture for multiple domains to synthesize realistic histological images while preserving the tissue structure.

#### Stain-aware models

Unlike the above stain normalization methods that project the external test data to the original training domain as a pre-processing step, stain information can be incorporated directly into the model, e.g., for nuclei segmentation by creating a hematoxylin-aware CNN (Zhao et al., 2020a). This approach is based on a U-Net (Ronneberger et al., 2015) and has three branches: one for processing the input image, one for processing the hematoxylin component (retrieved from a standard color deconvolution method), and one for feature aggregation of the other branches to finally output segmentation maps.

### 2.2 Tissue Type Segmentation

Accurate segmentation of a WSI into tissue types (e.g., epithelial vs. stromal vs. lymphatic tissue) allows for quantitative follow-up analysis. Depending on the kind of available annotations, we discriminate between the following categories: Methods for pixel-wise or patch-wise segmentation, hierarchical architectures that imitate a pathologist’s workflow, and methods that use WSI-level annotations and are therefore weakly supervised.

#### Pixel-wise segmentation

Vellal et al. (2021) assessed the risk of breast cancer from image features, such as the percentage of fibrous stroma, epithelium, and fatty tissue. To extract these features, they trained a 21-layer convolutional network inspired by VGG (Simonyan and Zisserman, 2014) and U-Net (Ronneberger et al., 2015) for pixel-wise segmentation of large 2048×2048 pixel tiles. Graham et al. (2020) developed a rotation-invariant CNN to account for the inherent rotational symmetry of histology images and validated their application on pixel-wise gland segmentation. Jayapandian et al. (2021) used pixel-wise segmentation for identifying six tissue types in kidney biopsies, applying the same U-Net architecture for segmenting patches with different magnifications.

#### Patch-wise segmentation

Image patches can be classified separately, and subsequently stitched together to create a coarse segmentation map of the entire WSI. From such segmented patches, Zhao et al. (2020b) computed a WSI’s tumor-stroma-ratio (TSR), a prognostic factor for colorectal cancer. Here, a pretrained VGG (Simonyan and Zisserman, 2014) was fine-tuned for classifying tiles into nine tissue types to determine the TSR.

Rączkowski et al. (2019) proposed an active learning framework to train a CNN, inspired by ResNet (He et al., 2016) and DarkNet (Redmon and Farhadi, 2017) for patch classification in colorectal cancer. The network’s uncertainty, estimated via Monte-Carlo dropout sampling (Gal and Ghahramani, 2015), was used to detect outlier tiles in the training set and to select them afterward for reconsideration. Wang et al. (2019) generated spatial tissue maps by classifying single cells into tumor, stroma, and lymphocytes. For this, they first extracted nuclei positions. Then, small patches centered around these nuclei were extracted and classified into the three classes using a CNN. The resulting classified positions can be used to extract spatial statistics or to generate segmentation masks using a kernel smoothing algorithm.

**Hierarchical segmentation** approaches mimic the workflow of pathologists by aggregating information from multiple scales of magnification. Schmitz et al. (2021) created a family of U-Net-based encoder-decoder architectures that process high- and low-resolution image tiles in separate branches from three publicly available datasets with liver, breast, and lymph node tissue. Additionally, they proposed a gate that decides whether to include the global context optimized by a classification loss to ease gradient flow through the deeper layers of the encoder for the global context. Alternatively, HookNet (van Rijthoven et al., 2021) fused the hidden space of multiple U-Net-based models that operate on different scales to deal with high resolution and contextual information in breast and lung cancer.

**Weakly supervised methods** typically require slide-level annotations only. One recent approach was training CNNs directly on the entire WSI of lung cancer sections (Chen et al., 2021). Subsequently, class activation maps (Zhou et al., 2016) highlight relevant cancerous regions that were also identified by pathologists, which can be interpreted as a confidence measure of the algorithm. Similarly, Silva-Rodríguez et al. (2021) trained a feature extraction network on the entire, downsampled WSI for the classification of global Gleason grades on prostate cancer. During inference, semantic segmentations are upscaled from the feature maps and achieve similar performance as fully supervised approaches while only using the global labels. Alternatively, the WSI can be split into patches, and patch-wise features used for WSI classification (Lu et al., 2021). Using this approach, attention scores produce interpretable heatmaps to visualize which regions contribute to the network’s prediction.

### 2.3 Evaluation of cell-level features

The evaluation of cell-level properties is a standard task in histopathology. For example, cell density and the abundance of dividing cells in tumor tissue are two important features for tumor grading. A multitude of deep learning approaches has been proposed for these and related tasks (Lagree et al., 2021). Here we focus on two of the most widely studied cell-level tasks, namely segmentation of nuclei and detection of mitoses.

#### Nuclei segmentation

To benchmark efforts in this field, segmentation challenges have been introduced, like the Multi-Organ Nucleus Segmentation Challenge (Kumar et al., 2020), which provided 30 images and around 22,000 nuclear boundary annotations in a public dataset. Out of the top six participants, three used U-Net-based semantic segmentation (Ronneberger et al., 2015), two used Mask R-CNN-based instance segmentation (He et al., 2017), and one group used stacked U-Net and R-CNN models. The two dominating algorithms have been further tailored towards nuclear segmentation: Cui et al. (2019) predicted a boundary map additionally to object segmentations to separate touching nuclei efficiently. Jin et al. (2020) incorporated a U-Net into a pipeline to detect lymph node metastasis in breast cancer patients, integrating multiple segmentation channels for nuclei, mitosis, tubule, etc. in the U-Net input. The Mask R-CNN has successfully been combined with a deep convolutional gaussian mixture color normalization model, which clusters pixels according to nucleus morphology (Jung et al., 2019), where the authors performed multiple interferences and post-processing steps to boost segmentation performance. Recently, other approaches such as GANs have been proposed (Mahmood et al., 2020a), where the network is trained on unpaired data to map segmentation masks to nuclei images. To ease the annotation process for nuclei segmentation, Qu et al. (2020) provided a deep learning framework trained on incomplete annotations, which are much easier to generate. Their two-stage approach first detected nuclei locations based on the partial annotations, before self-training with background propagation was applied to boost nuclei detection. In the second stage, a segmentation model was trained with this data in a weakly supervised fashion.

#### Mitosis detection

Identifying cells that are in the mitotic phases of the cell cycle is a diagnostically relevant task e.g. for breast cancer grading and prognosis (Veta et al., 2019). Several challenges released public datasets and benchmarked competing approaches for mitosis detection, e.g., ICPR MITOS-2012 (Roux et al., 2013), ICPR MITOS-ATYPIA-2014 (Roux et al., 2014), or TUPAC 16 (Veta et al., 2019). Most of the recently developed mitosis detection methods can be grouped into three categories: classification, segmentation, and detection. As one of the first deep learning applications in the medical field, Ciresan et al. (2013) trained a network to classify each pixel in an image by considering a tile centered around that pixel whilst tiles are extracted in a sliding-window fashion. This process was accelerated by precomputing the hematoxylin fraction of the H&E staining, which highlighted nuclei as mitosis candidates and hence restricted tile extraction (Saha et al., 2018). More recently, Pati et al. (2021) combined a classification task with metric learning to reduce the necessary amount of labeled data for more efficient network training. Another approach for mitosis detection is pixel-wise semantic segmentation. Jiménez and Racoceanu (2019) showed that a U-net-based semantic segmentation approach led to higher accuracy than previous classification approaches. Lafarge et al. (2021) proposed a special Euclidean motion group convolution to achieve translation and rotation invariance, which was integrated into a U-net architecture and improved the model’s robustness. Many other recent papers on mitosis detection were based on object detection (Lei et al., 2019, 2021; Sohail et al., 2021; Wollmann and Rohr, 2021), where only a weak centroid annotation that marks the center of the mitotic figure is required, compared to pixel-wise annotations for segmentation approaches. An alternative approach applied a cascade network, combining a first-stage object detection to identify mitosis candidates and a second-stage classification network for refinement (Mahmood et al., 2020b).

### 2.4 Genetic alteration prediction

As genetic alterations can carry crucial predictive and prognostic information, they have become increasingly relevant to the diagnostic workup and the selection of therapeutic pathways (Ashley, 2016). Therefore, patients are profiled for genetic alterations that characterize their disease, e.g., in colorectal cancer (Singh et al., 2021), to obtain better targeted therapies. However, using molecular assays to determine the mutational spectrum of malignant cells is expensive and time-consuming. Furthermore, DNA or RNA extracted from small samples may not suffice quantitatively for a comprehensive analysis, and RNA in older samples may already be degraded and hence not qualified for analysis. Techniques such as whole-genome sequencing require fresh tissue and are thus not applicable on formalin-fixed paraffin-embedded tissue that is usually used for histological sample preparation. Therefore, algorithm-based prognostic stratification and mutation prediction from H&E-stained WSIs offers an attractive, cost- and time-effective as well as tissue-sparing addition to existing molecular characterization methods.

#### Image-based mutation prediction

While some genetic alterations, such as mutations, copy number variations, and translocations can be relevant for disease characterization, the majority of work has so far focussed on mutation prediction from imaging data. Coudray et al. (2018) were the first to address this topic. They found that six commonly mutated genes in lung adenocarcinoma (STK11, EGFR, FAT1, SETBP1, KRAS, and TP53) can be predicted from WSI images. Since then, image-based mutation prediction has been applied to various types of cancers, such as melanoma (Kim et al., 2019; Zhang et al., 2020b), breast cancer (Anand et al., 2020; Bychkov et al., 2021; Lu et al., 2020), lung cancer (Wang et al., 2020; Yu et al., 2020), colorectal cancer (Cao et al., 2020; Echle et al., 2020b; Jang et al., 2020), bladder cancer (Woerl et al., 2020), and thyroid carcinoma (Tsou and Wu, 2019). Several recent studies attempt a pan-cancer approach that predicts genetic alteration across multiple tissue types from WSIs directly (Kather et al., 2020; Noorbakhsh et al., 2020).

#### Modeling strategies for mutation prediction

Most approaches rely on standardized processing pipelines from pre-processing (see Section 2.1) and region of interest extraction (Section 2.2) to model training and evaluation. As network architectures, common CNN models such as Inception (Szegedy et al., 2016) or ResNet (He et al., 2016) are used for per-tile prediction, where all tiles from a patient WSI inherit the same label. This label can be both continuous (e.g., tumor mutational burden) or categorical (e.g., the mutation status of a selected gene, or the microsatellite status) (Coudray et al., 2018; Kather et al., 2020). The final prediction at WSI level is an aggregation of tile labels using, in the simplest case, majority voting (for categorical targets) or averaging (for continuous targets). Alternatively, Cao et al. (2020) employ multiple instance learning (MIL) since it does not need instance labels for each tile and achieves better accuracy than standard supervised learning methods. Fu et al. (2020) did not train an end-to-end neural network for direct mutation prediction but rather classified each tile into different malignant and non-malignant tissue types. This classification network was used to extract features in a pan-cancer fashion and, subsequently, to predict driver gene mutations. Almost all studies mentioned above, except for Bychkov et al. (2021), trained their networks on publicly available data from The Cancer Genome Atlas (TCGA) (Gutman et al., 2013).

### 2.5 Grading, Staging, and Prognostic Information Extraction

A typical goal of the analysis of pathology slides is not only to recognize and evaluate primary lesions but also to determine their histopathologic subtype and grade (as defined by respective WHO classifications for different tumor entities) and to derive therapeutically relevant information from these features. In the context of computational pathology, this set of tasks can be addressed through the determination of features that are known to possess prognostic or predictive value. Alternatively, it can be attempted to extract prognostic or predictive information directly from imaging data, molecular properties, or clinical data. Both methodologies have recently been applied across a variety of entities.

#### Inference of known factors and biomarkers

Computational pathology approaches to extract known markers and scores include determination of the Gleason score in Prostate Cancer (Bulten et al., 2021; Nagpal et al., 2019; Steiner et al., 2020), grading of gliomas (Rathore et al., 2020; Truong et al., 2020), and automated evaluation of mitoses (Chang and Mrkonjic, 2020; Pantanowitz et al., 2020) or tumor-infiltrating lymphocytes in breast (Balkenhol et al., 2021) and head and neck cancer (Shaban et al., 2019a). Machine learning methods have also been applied in disease staging, e.g., to assess the degree of spread to the lymph nodes, either by highlighting areas suspicious for lymphovascular invasion as in the case of testicular cancer (Ghosh et al., 2021) or by predicting the risk of lymph node metastasis from the primary lesion in the case of bladder cancer (Harmon et al., 2020). Several studies inferred molecular properties with a prognostic value from H&E, such as microsatellite instability in gastrointestinal cancer (Kather et al., 2019a), or the molecular subtype of invasive bladder cancer (Woerl et al., 2020).

#### Image-based biomarkers

Finally, prognostic factors can be derived directly either from histopathology images (Zhao et al., 2020b) or in combination with other clinical or molecular data, e.g., from the genome or transcriptome (Failmezger et al., 2020; Hao et al., 2020; Zhao et al., 2020b). Potentially, this route can be followed without referring to previously known prognostic factors. Hence, while these approaches may first rely on computational predictions alone, they may also lead to the identification of novel prognostic or predictive factors that lend themselves to direct human evaluation, which can be identified through explainability methods (Tosun et al., 2020). Examples of this approach include automated quantification of intratumoral stroma in rectal cancer (Geessink et al., 2019), evaluation of nuclear morphology for survival prediction in lung cancer (Alsubaie et al., 2021), deep learning-based prognosis in nasopharyngeal cancer (Geessink et al., 2019; Zhang et al., 2020a), and survival prediction in colorectal cancer (Abbet et al., 2020; Kather et al., 2019b).

## 3 Methods

To assess reproducibility and reusability in computational pathology, we (i) monitored whether and how code was publicly available, (ii) evaluated criteria for data access, and (iii) checked if the statistical variance of the reported findings was provided. In Supplementary Table T2, we list all 42 publications (out of 161) together with the following evaluation criteria that we used for code, data, and statistical variance:

a. Inspired by the FAIR principles demanding that data should be findable, accessible, interoperable, and reusable (Wilkinson et al., 2016), we surveyed whether the code was made publicly available, in addition to also noting the platform that was used for sharing and the programming language and machine learning frameworks that were employed. Further, we checked for instructions for running the code, whether the code was minimally documented, and if a pretrained model was available for direct application.
b. For the 42 publications with available code, we evaluated data access and checked whether the dataset and required annotations were publicly available. Additionally, we recorded what kind of data had been used (e.g., tiled WSIs versus entire WSIs). We also reported whether the relevant pre-processing steps were provided as well as the training – validation – test split that was used for model development and evaluation. In terms of replicability, we specified what kind of test set had been used, whether it was similar to the training set or whether it covered an independent cohort.
c. Finally, we checked if any measure of statistical variance of the reported findings was provided. This is one way to tackle the difficulties concerned with reproducing the results, which can be introduced on multiple levels: computer-level inaccuracies like floating-point numbers that can be rounded differently on different machines, architectures, or execution environments (Hill, 2019), or algorithm-level stochasticity due to the stochastic behavior of optimization techniques. One way of dealing with this is to statistically analyze the experimental results and perform a sufficient exploration of hyperparameters (Pineau et al., 2020). A straightforward evaluation approach is to repeat the experiment multiple times and report mean and standard deviation across experiments, or over several folds of cross-validation.

## 4 Results

### Code availability

In our study, 42 out of 161 publications (26%) made their code publicly available (Fig. 3a). Interestingly, the ratio of publications with code differs across the five use cases: For stain normalization, we retrieved 29 research papers, where only 7 (24%) of them provided code with their method. In the field of tissue type segmentation and localization, only 12 out of our total 51 investigated papers (24%) had their code publicly available and only three publications provided the pretrained model weights. Among the 28 research papers that we screened for the evaluation of cell-level features, 11 papers (39%) provided code with the publications. The code of 5 of these 11 could be run in Google Colab and thus was directly applicable. For genetic alteration prediction, 8 out of 13 papers (61%) have provided their code along with their method. In survival analysis, only 4 out of 38 studies (11%) published code. Interestingly, genetic alteration prediction has the highest ratio of published code. One reason for this could be that a key publication for genetic alteration prediction published in Nature Medicine included a well-documented codebase (Coudray et al., 2018); most subsequent publications build on this work and could therefore publish their code more easily. Hence, the level of reusability in one field may depend on the preceding publications. This also strengthens the role of the publisher in the context of reproducibility and reusability in computational pathology. For the 42 papers that published their code, we checked the evaluation criteria for code, data, and statistical variance detailed in Section 3 and detailed them in Supplementary Table T2.

### Model weights and frameworks

Pretrained model weights were only available for half of the publications that also provided code (Fig. 3b); this renders a reproduction of experimental results difficult, in particular, because the pre-processing steps are rarely available. Also, without model weights a direct application without retraining the model is not possible, hence hampering its use by pathologists that are not specialized in deep learning. Almost 75% of the methods were implemented in Python using the open and publicly available machine learning frameworks TensorFlow^3^ (38%) and PyTorch^4^ (36%; Fig. 4c).

**Figure 4:**
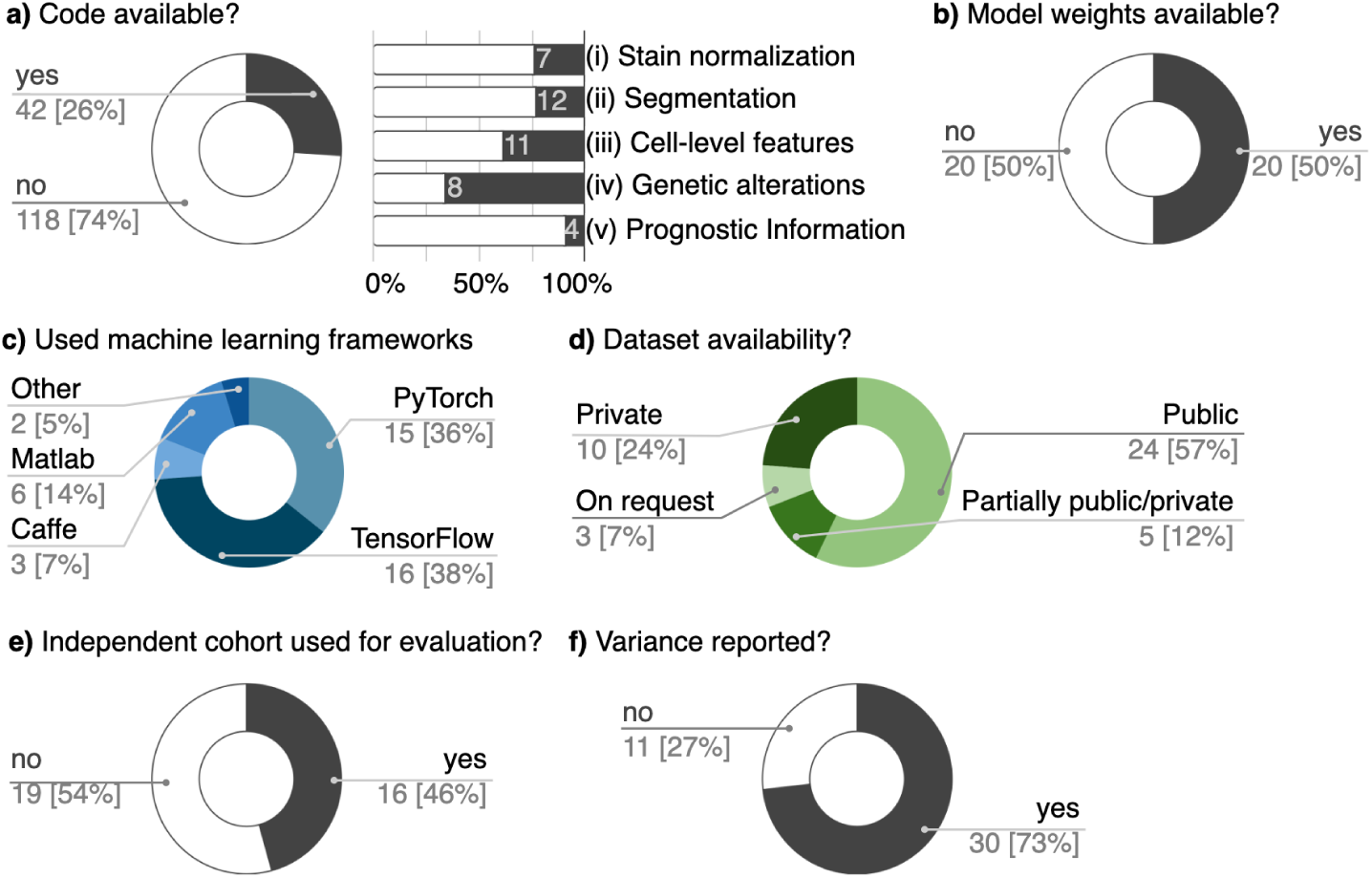
Analysis of our systematic literature search on 161 computational pathology papers. a) The proportion of methods with publicly available code (26%) differs across the use cases. b) Half of the publications with code release their final model weights. c) Most works (74%) used PyTorch or Tensorflow as machine learning frameworks. d) Mainly, large public datasets are used and sometimes complemented by private cohorts. e) Almost half of the publications used an independent cohort for evaluation. f) The largest part analyzes their results statistically.

### Datasets

More than half of all methods (57%) were evaluated on publicly available datasets (Fig. 3d). Most studies (e.g., all 13 studies reviewed for genetic alteration prediction) developed their methods based on TCGA (Gutman et al., 2013) it contains data from multiple institutions; thus, it can be split into training and test sets by cohort level. It was also a common practice to complement TCGA with external, mostly private data as an independent evaluation cohort. Nevertheless, TCGA, with a few hundred slides for each cancer type, is not sufficient to represent all cancer heterogeneity. The reliance of computational pathology on relatively few publicly available datasets renders their selection strategy and processing critical. Batch effects can be detected by deep learning models and lead to overestimation of the model’s performance (Howard et al., 2020). Therefore, we strongly encourage the development of more publicly available multi-institutional datasets.

### Statistical variance

We believe that a thorough evaluation of sources and magnitude of variability, both on an algorithmic and a data level, is an important step towards making modern computational pathology algorithms more reusable and generalizable. Almost three-quarters (73%) of the methods analyzed their results statistically, in which we considered all kinds of statistical notions to be statistical analysis (Fig. 3f). Many different sources of variability can be relevant to the performance of computational pathology algorithms, and it may therefore be difficult to devise a single strategy for quantifying all sources.

## 5 Conclusion

It has been increasingly recognized that computational reproducibility and reusability are an essential part of good scientific practice for deep learning applications (Haibe-Kains et al., 2020; Hutson, 2018; Stodden et al., 2016). Especially for the interdisciplinary field of computational pathology, both are key requirements for enabling a wider use of algorithms and eventually a clinical application.

In our survey of recently published computational pathology deep learning approaches however, we found that there is still a long way to go. For stain color normalization for example, techniques to reduce the color and intensity variations in histological images from different laboratories can render a downstream task algorithm more generalizable. Although neural network-based stain normalization techniques have evolved considerably in recent years (Section 2.1), their use in downstream applications is still limited, probably because pre-trained stain normalization models are rarely available and in most cases code is not shared. Instead, we observe that easy-to-use algorithms without further model training are typically applied. The lack of reusability hinders the practical application of innovative network-based methods.

Even if the code is shared, supporting documentation or convincing experiments on external cohorts are often missing, hence lowering the chances of a successful reuse and translation. Most state-of-the-art methods in computational pathology are based on deep learning algorithms, and typically require large amounts of labeled training data. Making this data available is as crucial as providing well documented code. We acknowledge that in some cases, data and appropriate annotations cannot be publicly shared, e.g. due to legal or ethical constraints. Here, reasonable compromises like partial data sharing or evaluation using public datasets (Haibe-Kains et al., 2020) should be considered.

Despite the lack of reproducibility and reusability in many computational pathology approches, we hope that the field will profit from the surging discussions, e.g. computer vision (Pineau et al., 2020). As a step in this direction, large conferences, such as MICCAI since 2021, started to employ reproducibility checklists for authors in their submission form that will be publicly available upon acceptance of the paper. We encourage the scientific community to recognize the long term value of reproducibility and reusability and to foster their realization in computational pathology.

## Data Availability

All data produced in the present work are contained in the manuscript.

## Author contributions

S.J.W., Ch.M., T.P., Ca.M. designed the concept of the review. M.B., Ch.M. supported with domain-related advice. S.J.W., L.L., D.W. compiled the evaluation criteria for reproducibility and reusability. S.J.W., Ch.M. assessed current data handling tools. For the use cases, S.J.W., D.W., S.S.B. reviewed the publications for stain normalization, L.L., S.J.W. for tissue type segmentation, D.W., T.P. for evaluation of cell-level features, T.P. for genetic alteration prediction, A.S., Ch.M. for grading, staging, and prognostic information extraction. S.J.W. analyzed the results. All authors contributed to constructive discussions. S.J.W. wrote the manuscript with Ch.M, Ca.M, T.P. and the input from all other authors.

## Acknowledgment

We thank Peter Schüffler (Munich) for inspiring feedback. S.J.W., L.L., and S.S.B. are supported by the Helmholtz Association under the joint research school “Munich School for Data Science - MUDS”. C.M. has received funding from the European Research Council (ERC) under the European Union’s Horizon 2020 research and innovation program (Grant agreement No. 866411)

## SUPPLEMENTARY INFORMATION

All publications listed in Table 0-5 were evaluated by our reproducibility criteria in July 2021.

### T0. Data Handling

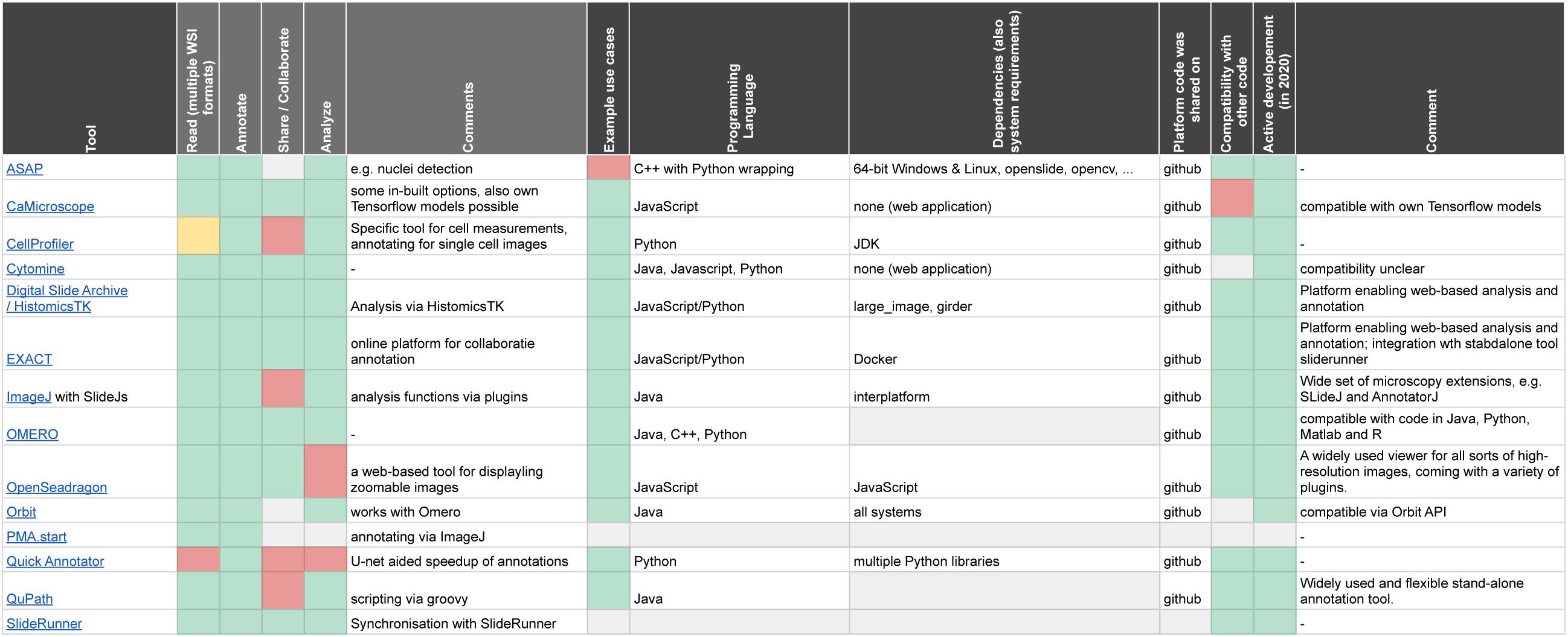

### T1. Stain normalization

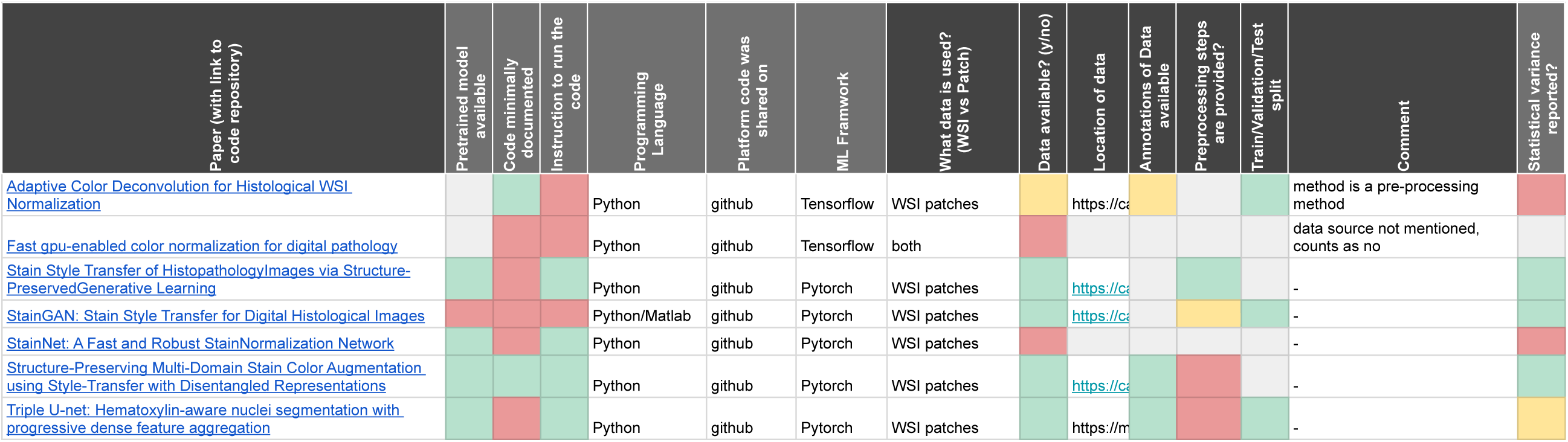

### T2. Tissue type segmentation

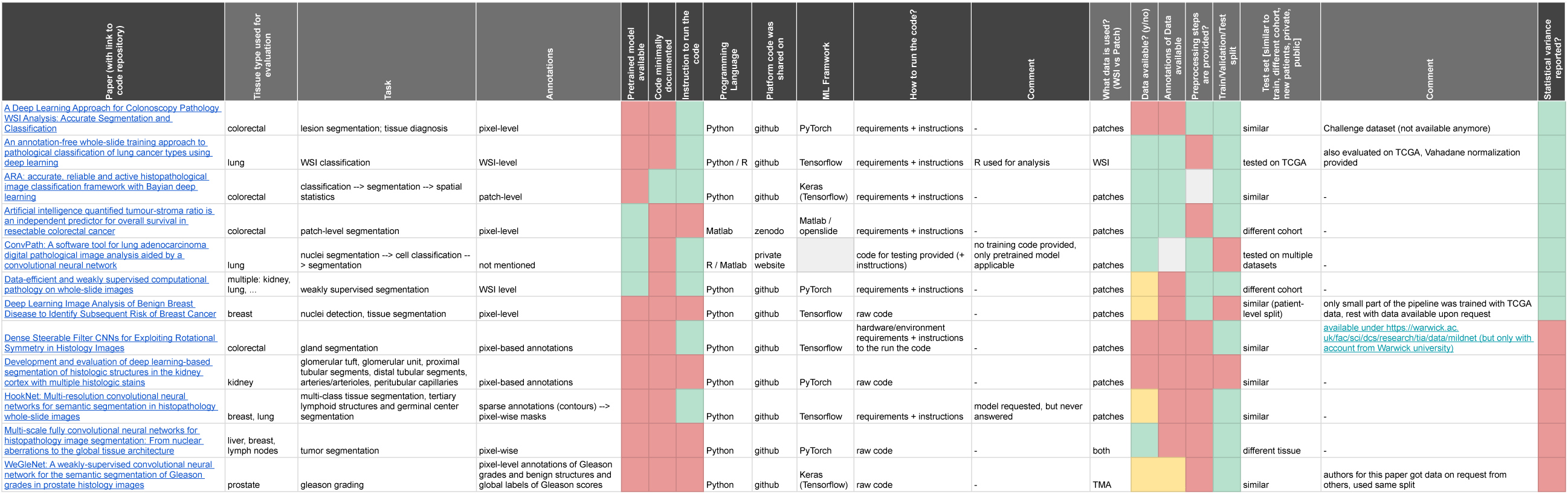

### T3. Evaluation of cell-level features

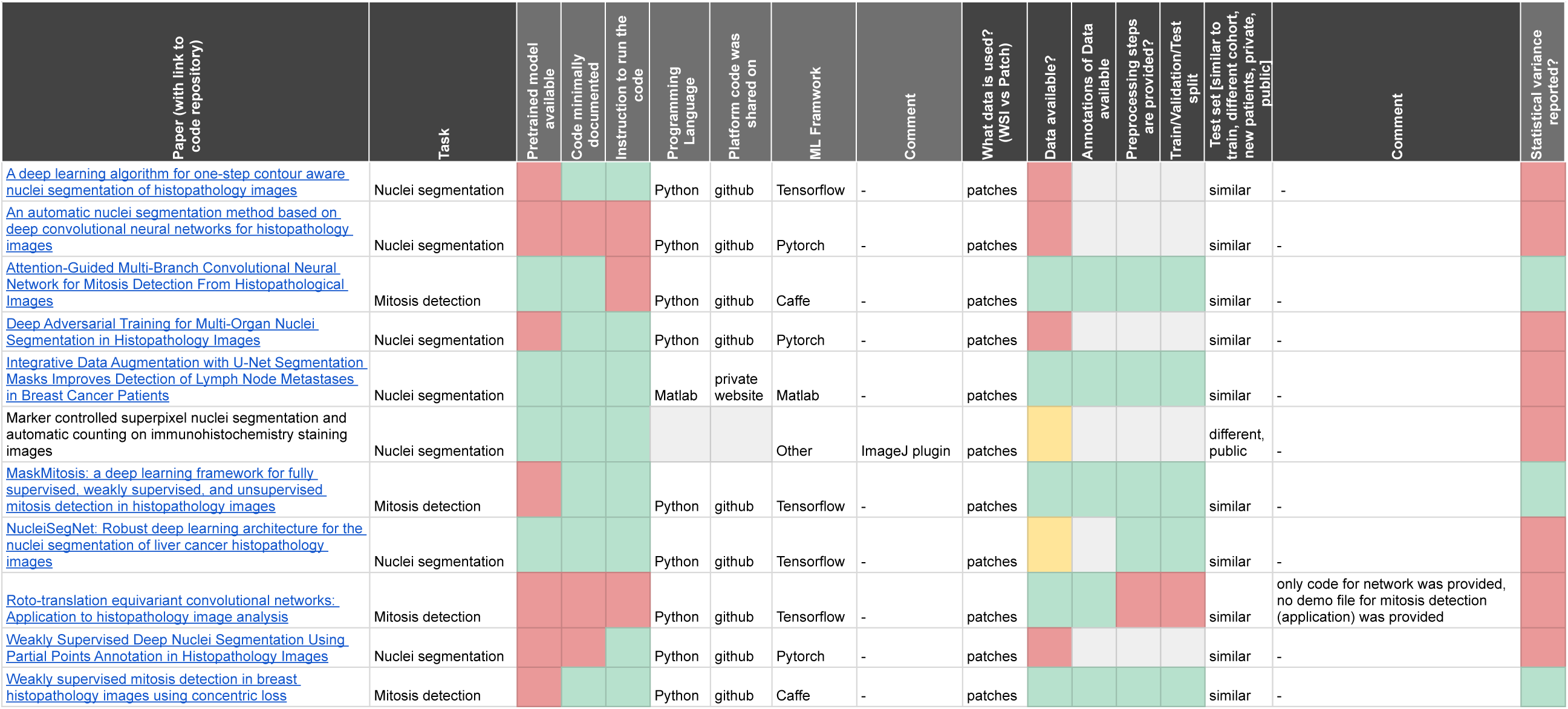

### T4. Genetic Alteration Prediction

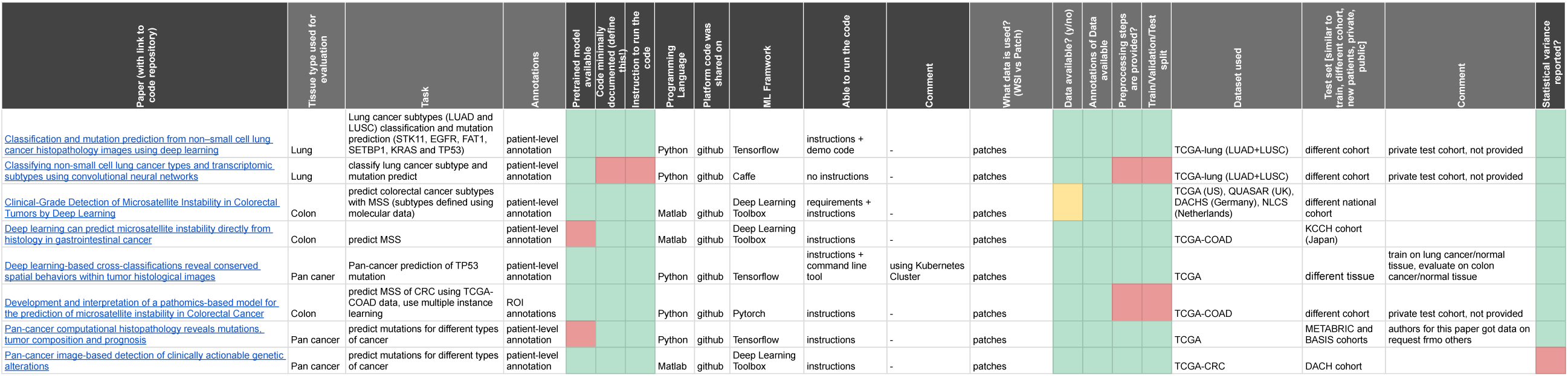

### T5. Grading, Staging, and Prognostic Information Extraction

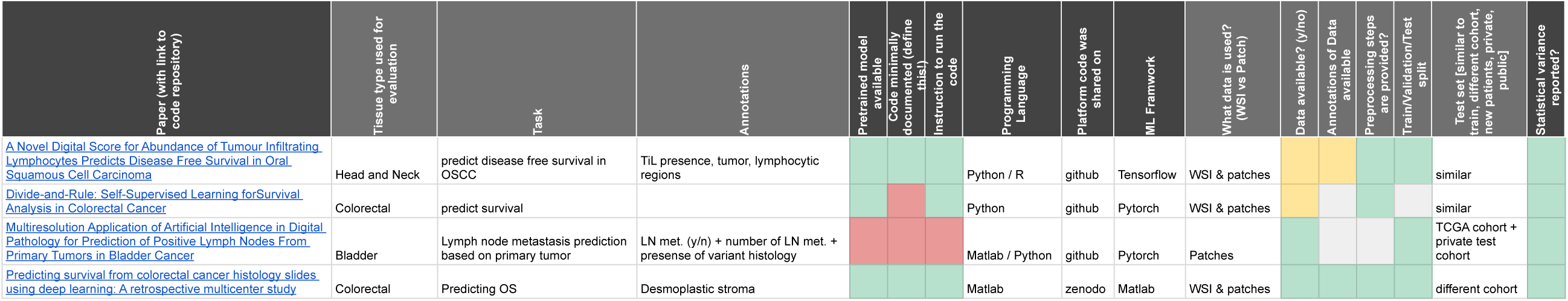

## Footnotes

1 https://computationalpathologygroup.github.io/ASAP/

2 https://github.com/camicroscope/caMicroscope

3 https://www.tensorflow.org/

4 https://pytorch.org/

## Notes

### Competing Interest Statement

The authors have declared no competing interest.

### Summary of Updates

corrected typos and layout

