## Supplementary Information for "Built to last? Reproducibility and Reusability of Deep Learning Algorithms in Computational Pathology" (from another version of this preprint: version 1, 2022-05-17)

All publications listed in Table 0-5 were evaluated by our reproducibility criteria in July 2021.

Legend:

- Yes
- No
- Partially / upon request
- Not applicable / not mentioned

| Tool | Read (multiple WSI formats) | Annotate | Share / Collaborate | Analyze | Comments | Example use cases | Programming Language | Dependencies (also system requirements) | Platform code was shared on | Compatibility with other code | Active developement (in 2020) | Comment |
|---|---|---|---|---|---|---|---|---|---|---|---|---|
| ASAP | | | | | e.g. nuclei detection | | C++ with Python wrapping | 64-bit Windows & Linux, openslide, opencv, ... | github | | | - |
| CaMicroscope | | | | | some in-built options, also own Tensorflow models possible | | JavaScript | none (web application) | github | | | compatible with own Tensorflow models |
| CellProfiler | | | | | Specific tool for cell measurements, annotating for single cell images | | Python | JDK | github | | | - |
| Cytomine | | | | | - | | Java, Javascript, Python | none (web application) | github | | | compatibility unclear |
| Digital Slide Archive / HistomicsTK | | | | | Analysis via HistomicsTK | | JavaScript/Python | large_image, girder | github | | | Platform enabling web-based analysis and annotation |
| EXACT | | | | | online platform for collaboratie annotation | | JavaScript/Python | Docker | github | | | Platform enabling web-based analysis and annotation; integration wth stabdalone tool sliderunner |
| ImageJ with SlideJs | | | | | analysis functions via plugins | | Java | interplatform | github | | | Wide set of microscopy extensions, e.g. SLideJ and AnnotatorJ |
| OMERO | | | | | - | | Java, C++, Python | | github | | | compatible with code in Java, Python, Matlab and R |
| OpenSeadragon | | | | | a web-based tool for displayling zoomable images | | JavaScript | JavaScript | github | | | A widely used viewer for all sorts of high-resolution images, coming with a variety of plugins. |
| Orbit | | | | | works with Omero | | Java | all systems | github | | | compatible via Orbit API |
| PMA.start | | | | | annotating via ImageJ | | | | | | | - |
| Quick Annotator | | | | | U-net aided speedup of annotations | | Python | multiple Python libraries | github | | | - |
| QuPath | | | | | scripting via groovy | | Java | | github | | | Widely used and flexible stand-alone annotation tool. |
| SlideRunner | | | | | Synchronisation with SlideRunner | | | | | | | - |

### T1. Stain normalization

| Paper (with link to code repository) | Pretrained model available | Code minimally documented | Instruction to run the code | Programming Language | Platform code was shared on | ML Framwork | What data is used? (WSI vs Patch) | Data available? (y/no) | Location of data | Annotations of Data available | Preprocessing steps are provided? | Train/Validation/Test split | Comment | Statistical variance reported? |
|---|---|---|---|---|---|---|---|---|---|---|---|---|---|---|
| Adaptive Color Deconvolution for Histological WSI Normalization | | | | Python | github | Tensorflow | WSI patches | | https://ca | | | | method is a pre-processing method | |
| Fast gpu-enabled color normalization for digital pathology | | | | Python | github | Tensorflow | both | | | | | | data source not mentioned, counts as no | |
| Stain Style Transfer of HistopathologyImages via Structure-PreservedGenerative Learning | | | | Python | github | Pytorch | WSI patches | | https://ca | | | | - | |
| StainGAN: Stain Style Transfer for Digital Histological Images | | | | Python/Matlab | github | Pytorch | WSI patches | | https://ca | | | | - | |
| StainNet: A Fast and Robust StainNormalization Network | | | | Python | github | Pytorch | WSI patches | | | | | | - | |
| Structure-Preserving Multi-Domain Stain Color Augmentation using Style-Transfer with Disentangled Representations | | | | Python | github | Pytorch | WSI patches | | https://ca | | | | - | |
| Triple U-net: Hematoxylin-aware nuclei segmentation with progressive dense feature aggregation | | | | Python | github | Pytorch | WSI patches | | https://m | | | | - | |

### T2. Tissue type segmentation

| Paper (with link to code repository) | Tissue type used for evaluation | Task | Annotations | Pretrained model available | Code minimally documented | Instruction to run the code | Programming Language | Platform code was shared on | ML Framwork | How to run the code? | Comment | What data is used? (WSI vs Patch) | Data available? (y/no) | Annotations of Data available | Preprocessing steps are provided? | Train/Validation/Test split | Test set [similar to train, different cohort, new patients, private, public] | Comment | Statistical variance reported? |
|---|---|---|---|---|---|---|---|---|---|---|---|---|---|---|---|---|---|---|---|
| A Deep Learning Approach for Colonoscopy Pathology WSI Analysis: Accurate Segmentation and Classification | colorectal | lesion segmentation; tissue diagnosis | pixel-level | | | | Python | github | PyTorch | requirements + instructions | - | patches | | | | | similar | Challenge dataset (not available anymore) | |
| An annotation-free whole-slide training approach to pathological classification of lung cancer types using deep learning | lung | WSI classification | WSI-level | | | | Python / R | github | Tensorflow | requirements + instructions | R used for analysis | WSI | | | | | tested on TCGA | also evaluated on TCGA, Vahadane normalization provided | |
| ARA: accurate, reliable and active histopathological image classification framework with Bayian deep learning | colorectal | classification --> segmentation --> spatial statistics | patch-level | | | | Python | github | Keras (Tensorflow) | requirements + instructions | - | patches | | | | | similar | - | |
| Artificial intelligence quantified tumour-stroma ratio is an independent predictor for overall survival in resectable colorectal cancer | colorectal | patch-level segmentation | pixel-level | | | | Matlab | zenodo | Matlab / openslide | requirements + instructions | - | patches | | | | | different cohort | - | |
| ConvPath: A software tool for lung adenocarcinoma digital pathological image analysis aided by a convolutional neural network | lung | nuclei segmentation --> cell classification --> segmentation | not mentioned | | | | R / Matlab | private website | | code for testing provided (+ insttructions) | no training code provided, only pretrained model applicable | patches | | | | | tested on multiple datasets | - | |
| Data-efficient and weakly supervised computational pathology on whole-slide images | multiple: kidney, lung, ... | weakly supervised segmentation | WSI level | | | | Python | github | PyTorch | requirements + instructions | - | patches | | | | | different cohort | - | |
| Deep Learning Image Analysis of Benign Breast Disease to Identify Subsequent Risk of Breast Cancer | breast | nuclei detection, tissue segmentation | pixel-level | | | | Python | github | Tensorflow | raw code | - | patches | | | | | similar (patient-level split) | only small part of the pipeline was trained with TCGA data, rest with data available upon request | |
| Dense Steerable Filter CNNs for Exploiting Rotational Symmetry in Histology Images | colorectal | gland segmentation | pixel-based annotations | | | | Python | github | Tensorflow | hardware/environment requirements + instructions to the run the code | - | patches | | | | | similar | available under https://warwick.ac.uk/fac/sci/dcs/research/tia/data/mildnet (but only with account from Warwick university) | |
| Development and evaluation of deep learning-based segmentation of histologic structures in the kidney cortex with multiple histologic stains | kidney | glomerular tuft, glomerular unit, proximal tubular segments, distal tubular segments, arteries/arterioles, peritubular capillaries | pixel-based annotations | | | | Python | github | PyTorch | raw code | - | patches | | | | | similar | - | |
| HookNet: Multi-resolution convolutional neural networks for semantic segmentation in histopathology whole-slide images | breast, lung | multi-class tissue segmentation, tertiary lymphoid structures and germinal center segmentation | sparse annotations (contours) --> pixel-wise masks | | | | Python | github | Tensorflow | requirements + instructions | model requested, but never answered | patches | | | | | similar | - | |
| Multi-scale fully convolutional neural networks for histopathology image segmentation: From nuclear aberrations to the global tissue architecture | liver, breast, lymph nodes | tumor segmentation | pixel-wise | | | | Python | github | PyTorch | raw code | - | both | | | | | different tissue | - | |
| WeGleNet: A weakly-supervised convolutional neural network for the semantic segmentation of Gleason grades in prostate histology images | prostate | gleason grading | pixel-level annotations of Gleason grades and benign structures and global labels of Gleason scores | | | | Python | github | Keras (Tensorflow) | raw code | - | TMA | | | | | similar | authors for this paper got data on request from others, used same split | |

### T3. Evaluation of cell-level features

| Paper (with link to code repository) | Task | Pretrained model available | Code minimally documented | Instruction to run the code | Programming Language | Platform code was shared on | ML Framwork | Comment | What data is used? (WSI vs Patch) | Data available? | Annotations of Data available | Preprocessing steps are provided? | Train/Validation/Test split | Test set [similar to train, different cohort, new patients, private, public] | Comment | Statistical variance reported? |
|---|---|---|---|---|---|---|---|---|---|---|---|---|---|---|---|---|
| A deep learning algorithm for one-step contour aware nuclei segmentation of histopathology images | Nuclei segmentation | | | | Python | github | Tensorflow | - | patches | | | | | similar | - | |
| An automatic nuclei segmentation method based on deep convolutional neural networks for histopathology images | Nuclei segmentation | | | | Python | github | Pytorch | - | patches | | | | | similar | - | |
| Attention-Guided Multi-Branch Convolutional Neural Network for Mitosis Detection From Histopathological Images | Mitosis detection | | | | Python | github | Caffe | - | patches | | | | | similar | - | |
| Deep Adversarial Training for Multi-Organ Nuclei Segmentation in Histopathology Images | Nuclei segmentation | | | | Python | github | Pytorch | - | patches | | | | | similar | - | |
| Integrative Data Augmentation with U-Net Segmentation Masks Improves Detection of Lymph Node Metastases in Breast Cancer Patients | Nuclei segmentation | | | | Matlab | private website | Matlab | - | patches | | | | | similar | - | |
| Marker controlled superpixel nuclei segmentation and automatic counting on immunohistochemistry staining images | Nuclei segmentation | | | | | | Other | ImageJ plugin | patches | | | | | different, public | - | |
| MaskMitosis: a deep learning framework for fully supervised, weakly supervised, and unsupervised mitosis detection in histopathology images | Mitosis detection | | | | Python | github | Tensorflow | - | patches | | | | | similar | - | |
| NucleiSegNet: Robust deep learning architecture for the nuclei segmentation of liver cancer histopathology images | Nuclei segmentation | | | | Python | github | Tensorflow | - | patches | | | | | similar | - | |
| Roto-translation equivariant convolutional networks: Application to histopathology image analysis | Mitosis detection | | | | Python | github | Tensorflow | - | patches | | | | | similar | only code for network was provided, no demo file for mitosis detection (application) was provided | |
| Weakly Supervised Deep Nuclei Segmentation Using Partial Points Annotation in Histopathology Images | Nuclei segmentation | | | | Python | github | Pytorch | - | patches | | | | | similar | - | |
| Weakly supervised mitosis detection in breast histopathology images using concentric loss | Mitosis detection | | | | Python | github | Caffe | - | patches | | | | | similar | - | |

### T4. Genetic Alteration Prediction

| Paper (with link to code repository) | Tissue type used for evaluation | Task | Annotations | Pretrained model available | Code minimally documented (define this!) | Instruction to run the code | Programming Language | Platform code was shared on | ML Framwork | Able to run the code | Comment | What data is used? (WSI vs Patch) | Data available? (y/no) | Annotations of Data available | Preprocessing steps are provided? | Train/Validation/Test split | Dataset used | Test set [similar to train, different cohort, new patients, private, public] | Comment | Statistical variance reported? |
|---|---|---|---|---|---|---|---|---|---|---|---|---|---|---|---|---|---|---|---|---|
| Classification and mutation prediction from non–small cell lung cancer histopathology images using deep learning | Lung | Lung cancer subtypes (LUAD and LUSC) classification and mutation prediction (STK11, EGFR, FAT1, SETBP1, KRAS and TP53) | patient-level annotation | | | | Python | github | Tensorflow | instructions + demo code | - | patches | | | | | TCGA-lung (LUAD+LUSC) | different cohort | private test cohort, not provided | |
| Classifying non-small cell lung cancer types and transcriptomic subtypes using convolutional neural networks | Lung | classify lung cancer subtype and mutation predict | patient-level annotation | | | | Python | github | Caffe | no instructions | - | patches | | | | | TCGA-lung (LUAD+LUSC) | different cohort | private test cohort, not provided | |
| Clinical-Grade Detection of Microsatellite Instability in Colorectal Tumors by Deep Learning | Colon | predict colorectal cancer subtypes with MSS (subtypes defined using molecular data) | patient-level annotation | | | | Matlab | github | Deep Learning Toolbox | requirements + instructions | - | patches | | | | | TCGA (US), QUASAR (UK), DACHS (Germany), NLCS (Netherlands) | different national cohort | | |
| Deep learning can predict microsatellite instability directly from histology in gastrointestinal cancer | Colon | predict MSS | patient-level annotation | | | | Matlab | github | Deep Learning Toolbox | instructions | - | patches | | | | | TCGA-COAD | KCCH cohort (Japan) | | |
| Deep learning-based cross-classifications reveal conserved spatial behaviors within tumor histological images | Pan caner | Pan-cancer prediction of TP53 mutation | patient-level annotation | | | | Python | github | Tensorflow | instructions + command line tool | using Kubernetes Cluster | patches | | | | | TCGA | different tissue | train on lung cancer/normal tissue, evaluate on colon cancer/normal tissue | |
| Development and interpretation of a pathomics-based model for the prediction of microsatellite instability in Colorectal Cancer | Colon | predict MSS of CRC using TCGA-COAD data, use multiple instance learning | ROI annotations | | | | Python | github | Pytorch | instructions | - | patches | | | | | TCGA-COAD | different cohort | private test cohort, not provided | |
| Pan-cancer computational histopathology reveals mutations, tumor composition and prognosis | Pan cancer | predict mutations for different types of cancer | patient-level annotation | | | | Python | github | Tensorflow | instructions | - | patches | | | | | TCGA | METABRIC and BASIS cohorts | authors for this paper got data on request frmo others | |
| Pan-cancer image-based detection of clinically actionable genetic alterations | Pan cancer | predict mutations for different types of cancer | patient-level annotation | | | | Matlab | github | Deep Learning Toolbox | instructions | - | patches | | | | | TCGA-CRC | DACH cohort | | |

#### T5. Grading, Staging, and Prognostic Information Extraction

| Paper (with link to code repository) | Tissue type used for evaluation | Task | Annotations | Pretrained model available | Code minimally documented (define this!) | Instruction to run the code | Programming Language | Platform code was shared on | ML Framwork | What data is used? (WSI vs Patch) | Data available? (y/no) | Annotations of Data available | Preprocessing steps are provided? | Train/Validation/Test split | Test set [similar to train, different cohort, new patients, private, public] | Statistical variance reported? |
|---|---|---|---|---|---|---|---|---|---|---|---|---|---|---|---|---|
| A Novel Digital Score for Abundance of Tumour Infiltrating Lymphocytes Predicts Disease Free Survival in Oral Squamous Cell Carcinoma | Head and Neck | predict disease free survival in OSCC | TiL presence, tumor, lymphocytic regions | | | | Python / R | github | Tensorflow | WSI & patches | | | | | similar | |
| Divide-and-Rule: Self-Supervised Learning forSurvival Analysis in Colorectal Cancer | Colorectal | predict survival | | | | | Python | github | Pytorch | WSI & patches | | | | | similar | |
| Multiresolution Application of Artificial Intelligence in Digital Pathology for Prediction of Positive Lymph Nodes From Primary Tumors in Bladder Cancer | Bladder | Lymph node metastasis prediction based on primary tumor | LN met. (y/n) + number of LN met. + presense of variant histology | | | | Matlab / Python | github | Pytorch | Patches | | | | | TCGA cohort + private test cohort | |
| Predicting survival from colorectal cancer histology slides using deep learning: A retrospective multicenter study | Colorectal | Predicting OS | Desmoplastic stroma | | | | Matlab | zenodo | Matlab | WSI & patches | | | | | different cohort | |
